# ‘*That’s why I wanted him to go on dialysis*’ – a qualitative inductive thematic analysis of older patients’ and their family members’ perspectives on kidney failure treatment decision-making

**DOI:** 10.1101/2025.04.26.25326402

**Authors:** Robert A Kimmitt, Charlotte M Snead, Leila Rooshenas, Fergus J Caskey, Joanna Coast, Rachael L. Morton, Peter Quartermaine, Luisa Quartermaine, Lucy E. Selman, Barnaby Hole

## Abstract

**Background:** Chronic kidney disease (CKD) is increasingly common amongst frail older patients with multiple health problems. These patients typically decide between kidney replacement therapy (KRT) with dialysis, which carries uncertain survival benefit with significant treatment burden, and conservative kidney management (CKM). A person-centred approach to this shared decision-making process is advocated. Family members are known to be important in these decisions. Nonetheless, data exploring family member perspectives are limited. We aimed to explore how older, frail and/or comorbid patients with CKD and their family members understand and approach decision-making regarding treatments for kidney failure.

**Methods:** Semi-structured interviews were conducted, in person, in 2018-2019, with older patients with advanced CKD (≥80 years or ≥65 with evidence of frailty or comorbidity) and at least one family member per patient. Interview transcripts were analysed using inductive thematic analysis with constant comparison within and between family units. Meanings and concepts were discussed between study investigators, to generate a coding framework and develop major themes.

**Results:** Ten patients and 12 associated family members were interviewed. Three major themes were identified: (1) *“whose decision is it anyway?”* concerns ownership of treatment decisions; (2) *“on death, dying and uncertain futures”* describes relational elements of participants’ thoughts of the future; and (3) *“caring and being cared for”* explores the importance of physical and emotional caring roles and love and care in relationships.

**Conclusions:** Family members appear to have significant influence on older patients’ kidney failure treatment decisions, which can occur outside the shared decision-making support offered to patients. The inextricably intertwined daily lives of co-habiting patients and family members means that treatment decisions impact and depend upon both family members and patients. Kidney services should adopt a ‘family-centred’ (rather than individually ‘person-centred’) approach to decision-making support and must develop ways to embed this in clinical practice.

## Background

Chronic kidney disease (CKD) is increasingly common in the UK and globally, especially amongst older adults with multiple health problems.^1,2^ The median age of patients starting kidney replacement therapy (KRT) in the UK in 2021 was 63.7,^1^ and is higher in many other European countries.^3^ Meanwhile, the benefits of KRT over conservative kidney management (CKM) in older, frailer patients are unclear.^4–6^ Whilst observational evidence suggests a survival benefit associated with preparing for KRT, this may be lost in those who are over 80 years, or over 65 with multiple comorbidities.^4^ As such, older, frailer patients approaching kidney failure make decisions about their treatment in the face of uncertain potential benefits^7^ and potential disruption to both their own lives and the lives of those close to them, at a time when their capacity to deal with this decision may be low.^8^

Family members are known to contribute to decisions around treatments for kidney failure by expressing their values and opinions,^9–13^ providing social support,^9,14,15^ including for implementing home therapies,^12,14^ and by gaining and sharing knowledge of treatment options.^9,14^ Family members may also be receiving physical or emotional care from patients^10^ and it is known that patients can strive to avoid burdening those close to them.^9,16^ Much of this existing data comes from patients and clinicians, however, rather than directly from family members.^9^ Previous research about kidney failure decision-making involving patient-family member dyads has focused on dialysis and transplantation, and CKM has been under-represented.^17,18^ Work in older patients has identified the significant burdens on caregivers of patients receiving CKM, which are thought to be comparable to those of caregivers of patients receiving dialysis.^19,20^ These data are not dyadic and include patients (and caregivers) already committed to a treatment, not those making decisions about treatment options. There is consequently little data investigating relationships and dynamics between older patients and their family members considering CKM to inform clinicians supporting such decision-making.^9,13,21^

We aimed to explore how older, frail and/or comorbid patients with CKD and their family members understand and approach decision-making regarding treatments for kidney failure.

## Methods

### Study design

A social constructionist study design was used, involving inductive thematic analysis of qualitative interviews with family groups, each including an older patient with advanced CKD and at least one family member.

### Inclusion and exclusion criteria

#### Patient participants

Inclusion criteria:

– English-speaking

– Cared for by renal services with estimated glomerular filtration rate (eGFR) below 15ml/min/1.73m^2^ (on two blood tests in the previous 12 months)

– ≥80 years old, or ≥65 years old with a Davies comorbidity score of ≥2,^22^ and/or World Health Organization (WHO) performance status of ≥3^23^

Exclusion criteria:

– Previous outpatient KRT

– Due to start KRT within the next month

– Unable to consent

These criteria were designed to select patients for whom the benefits of dialysis versus CKM are not unequivocally established,^4,5^ as this work was undertaken as part of a larger study investigating treatment preferences of older patients considering CKM and dialysis.^24,25^

#### Family member participants

Inclusion criteria:

– English-speaking

– Aged 18 years or older

– Nominated by eligible patients as a close person with whom they would discuss treatment decisions.

Exclusion criterion:

– Unable to consent

The term “family members” is used throughout this paper to refer to people close to patients (including family, close friends and chosen family), and for clarity we use the term “patients” when describing people with kidney disease.

### Sampling and recruitment

Purposive sampling sought to optimise variety across clinical and demographic characteristics. Up to two family members were invited by the patient participant and asked to contact the study team if they were interested to take part. Analysis and recruitment were conducted in parallel and recruitment ceased at data saturation, i.e. when no new major themes were identified.^26^

Recruitment took place in three kidney units in England. Two were tertiary renal centres offering transplantation, dialysis and specialised CKM services, the third was a non-transplant centre with 200 dialysis recipients which delivered CKM under general nephrology services. Local NHS kidney teams (comprising specialist doctors and nurses) identified eligible participants in outpatient clinics and provided verbal information and written materials before the study team arranged interviews.

### Data collection

Semi-structured, face-to-face interviews were conducted with participants in their homes between September 2018 and July 2019 by BH – a white, cis-male doctor in his late 30s, training to be a kidney specialist. BH had received training in qualitative research and interviewing skills; this work was undertaken during his PhD and was his first experience of qualitative research. He was not known to the participants prior to the arrangement of the interviews, and described himself as a “researcher”, not disclosing his clinical role except on the one occasion where he was asked about this. Written consent was collected alongside demographic information. The interviews used open-ended questioning supported by a topic guide (supplementary figure 1) developed based on the literature and with input from patient and public involvement group members. Interviews were audio-recorded using an encrypted digital recorder, and field notes were taken to provide context for analysis. Recordings were transcribed word-for-word and participant names pseudonymised before analysis, to maintain anonymity.

### Ethical considerations

This research was conducted to a high ethical standard. Participants were recruited voluntarily and gave informed consent with the ability to opt out at any time without needing to give a reason. Efforts were made to ensure participant anonymity and confidentiality. A risk mitigation strategy was employed to manage to possibility of participants becoming distressed by discussions around sensitive topics of death and dying, familial conflict, and treatment uncertainty. This included a distress protocol which is included as supplementary figure 2.

The study was reviewed by Surrey Research Ethics Committee and granted NHS Health Research Authority and Health and Care Research Wales approval on 2/8/18 (ref: 18/LO/1179). Reporting is in accordance with the Consolidated Criteria for Reporting Qualitative research (COREQ)^27^ (supplementary table 1).

### Patient and public involvement (PPI)

PPI work throughout the research process is crucial for the delivery of high-quality qualitative research, because it increases the relevance of the research for patients and family members, helps to ensure safe and ethically conducted research, and can help with data interpretation to maximise validity.^28^ Patients and members of the public were involved in this research at every stage in its delivery, beginning in 2015 with two PPI group meetings which were integral to the conception of the study idea and design, including the interview topic guide, risk mitigation strategy and distress protocol. PPI group meetings and oversight continued throughout the interview stage of the research. The PPI work included a mixed patient/public conference event which included a play based on one of the interview transcripts. The original PPI group members were not available for the analysis stage but our patient and family member co-authors (PQ and LQ) helped with data interpretation (including multiple discussions around thematic development and the significance of thematic findings, and co-authoring the paper).

### Methodological approach and theoretical stance

The study adopts a constructionist theoretical stance with a critical realist ontology and subjectivist epistemology.^29^ Critical realism, originally developed by Bhaskar,^30^ posits deeper, underlying structures in the world that exist independently of our awareness, regardless of our ability to come to know them. Subjectivism is the belief that knowledge is “always filtered through the lenses of language, gender, social class, race, and ethnicity”^31(p.21)^ A subjectivist epistemology recognizes knowledge as value laden.

### Analysis

Interviews where a patient and at least one family member (dyads or triads) took part were included in this analysis, and were analysed using inductive thematic analysis. Coding was primarily inductive and used constant comparison.^32,33^ Thematic analysis followed the 6 steps described by Braun and Clarke: 1) familiarisation with the data; 2) initial coding; 3) searching for themes; 4) reviewing themes; 5) defining themes; and 6) producing the report.^32^ After familiarisation with the dataset, RK and CS independently line-by-line coded all of the transcripts. BH line-by-line coded all of the patient transcripts and the first three family member transcripts before reviewing RK’s and CS’s line-by-line coding of all of the family member transcripts. BH led on the primary analysis of the patient data^25^ while RK led on this analysis of the family member data. RK, BH and CS met to discuss initial themes, concepts and meanings. Analytic accounts were first written (using Microsoft Word) by RK for each family unit. Family unit analytical accounts were then progressively amalgamated (with ongoing constant comparison within and between dyads/triads) towards a master analytic account for all participants. A thematic framework was developed from this data, with input from all co-authors, focussing on the impact of family members and the interplay between patients and family members in treatment decision-making. PQ and LQ were involved as patient and family member co-authors, contributing to critical analysis and data interpretation once the master analytic account had been developed.

### Rigour and reflexivity

Rigour was a key consideration throughout the planning, interview and analysis stages of this work. BH kept detailed field notes and wrote reflections after line-by-line coding the patient participants. Team meetings including discussions about reflexivity and RK kept a reflexivity diary during the analysis and writing stage. A clear audit trail was kept documenting the different stages in the analysis which are summarised above. The double-or triple-coding of interview data (with detailed discussion between coders) and the reproduction of a wide range of primary data excerpts was used to further support rigour.

## Results

33 patients were approached, of whom 10 were interviewed and recruited one or more family members. 5 patients were interviewed but did not recruit a family member, so were not included in this dyadic analysis (but these interview data contributed to other analyses within the wider research project)^24,25^. 18 patients were approached but declined to take part, with reasons including: being too busy [2], memory problems [1], deafness [2], being away [1], having started dialysis [1] and being in hospital [2] (supplementary figure 3). Data from 10 patients and 12 family members (22 participants in total) were therefore included in this analysis (table 1).

**Table 1:** Participant characteristics.

| Patients (n=10) |  | Family members (n=12) |  |
| --- | --- | --- | --- |
| **Gender:** |  | **Gender:** |  |
| - Male | 6 (60%) | - Male | 3 (25%) |
| - Female | 4 (40%) | - Female | 9 (75%) |
| **Age:** |  | **Relationship to patient:** |  |
| - 65-69 | 1 (10%) | - Spouse/partner | 6 (50%) |
| - 70-74 | 0 | - Adult child | 5 (42%) |
| - 75-79 | 2 (20%) | - Adult grandchild | 1 (8%) |
| - 80-84 | 3 (30%) |  |  |
| - 85-89 | 3 (30%) |  |  |
| - 90-94 | 1 (10%) |  |  |
| **eGFR (ml/min/1.73m<sup>2</sup>):** |  | **Age:** |  |
| - <10 | 3 (30%) | - 40-44 | 2 (17%) |
| - 10-14 | 7 (70%) | - 45-49 | 0 |
| **Treatment plan:** |  | - 50-54 | 2 (17%) |
| - HD | 4 (40%) | - 55-59 | 0 |
| - PD | 1 (10%) | - 60-64 | 2 (17%) |
| - CKM | 5 (50%) | - 65-69 | 1 (8%) |
| - Listed for transplantation | 0 | - 70-74 | 2 (17%) |
| **Major comorbidities:** |  | - 75-79 | 2 (17%) |
| - Type 2 diabetes | 7 (70%) | - 80-84 | 1 (8%) |
| - Ischaemic heart disease | 7 (70%) |  |  |
| - Hypertension | 3 (30%) |  |  |
| - Malignancy | 4 (40%) |  |  |
| - Obesity | 2 (20%) |  |  |
| - Heart failure | 1 (10%) |  |  |
| - Other comorbidity | 3 (30%) |  |  |
| **WHO performance status:** |  |  |  |
| - 0 | 0 |  |  |
| - 1 | 3 (30%) |  |  |
| - 2 | 2 (20%) |  |  |
| - 3 | 5 (50%) |  |  |
| - 4 | 0 |  |  |
| **Years of FTE:** |  |  |  |
| - 0-5 | 0 |  |  |
| - 6-10 | 5 (50%) |  |  |
| - 11-15 | 4 (40%) |  |  |
| - 16-20 | 1 (10%) |  |  |
| **Family members recruited:** |  |  |  |
| - 1 | 8 (80%) |  |  |
| - 2 | 2 (20%) |  |  |

eGFR: estimated glomerular filtration rate, HD: haemodialysis, PD: peritoneal dialysis, CKM: conservative kidney management, WHO: World Health Organisation, FTE: full time education

Patient ages were 65-90 years, six (60%) were male, all were retired, eGFR ranged from 7-14mL/min/1.73m^2^, all described their ethnicity as white British. Diabetes was the most common reason for CKD. Five patients were planning for CKM, four for haemodialysis (HD) and one for peritoneal dialysis (PD), with none on a transplantation waiting list (though transplant ‘eligibility’ was neither an inclusion or exclusion criterion). Family members comprised six partners/spouses, five adult children, and one adult grandchild (table 1).

Patient interviews lasted on average 57 minutes (range 29-81) and family member interviews lasted on average 51 minutes (range 31-80). Three key themes illustrating the importance of family members for patients’ decision-making processes were identified. “*Whose decision is it anyway?*” speaks to variations and nuances around the ownership of kidney failure treatment decisions. “*On death, dying and uncertain futures*” describes participants’ thoughts of the future. “*Caring and being cared for*” explores the importance of physical and emotional caring roles and the importance of love and care in relationships. Table 2 provides illustrative quotations for each theme.

**Table 2:**
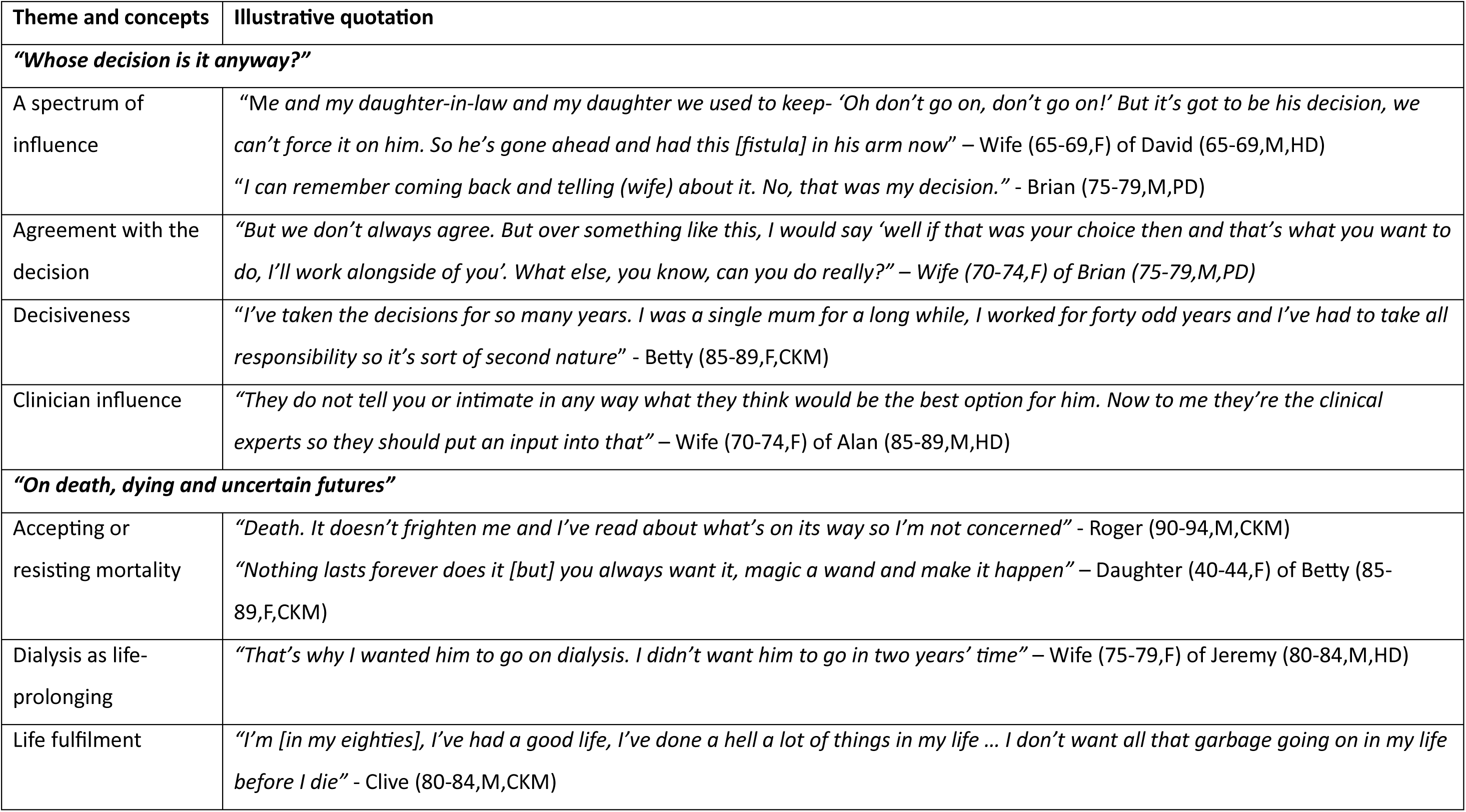
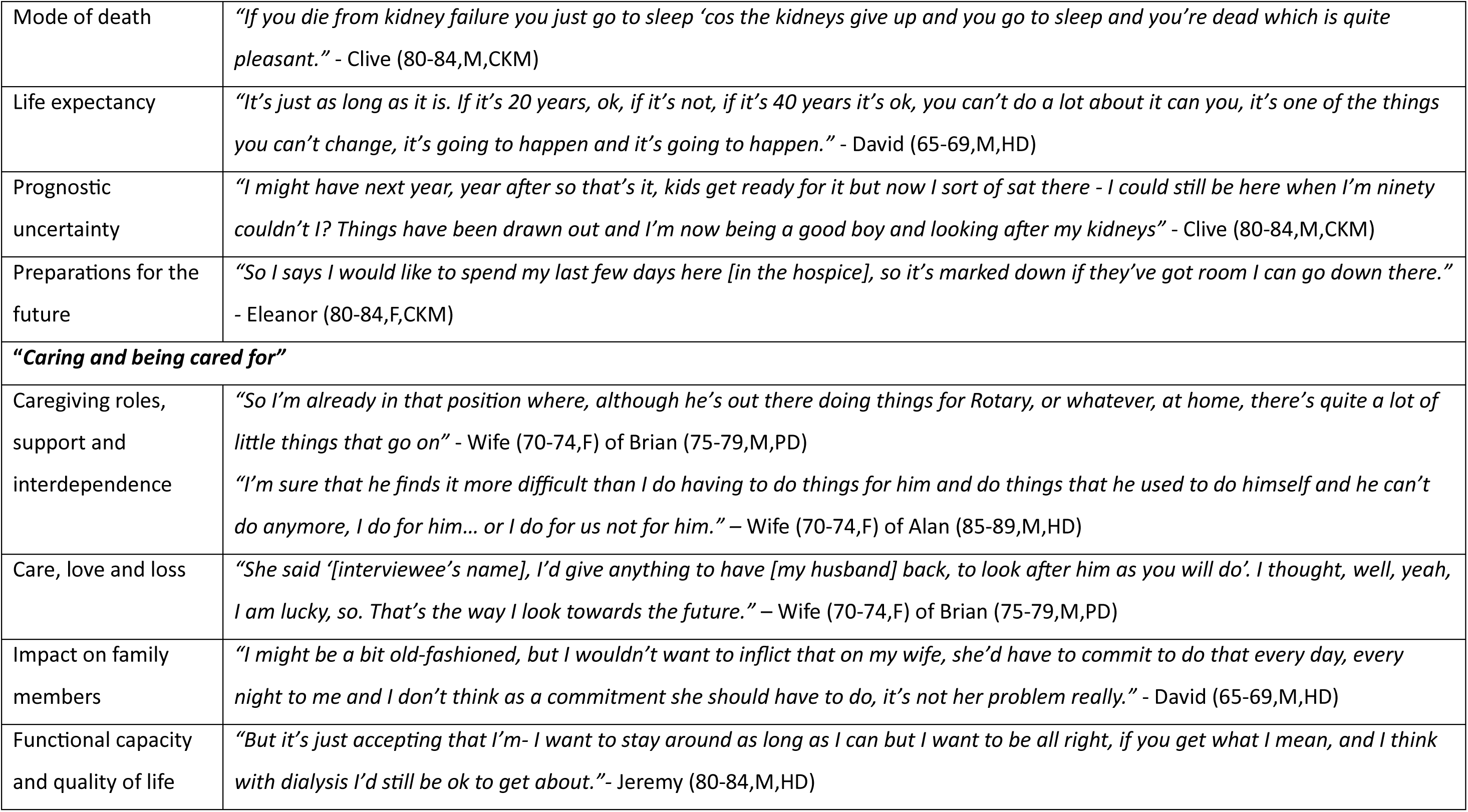
Illustrative quotations for themes and concepts.

Exemplifying data extracts are included to demonstrate analytical points, aiming to feature a variety of participants and represent deviant cases. Quotations are marked with patients’ pseudonym, age (category), gender (M for male, F for female) and documented treatment plan (HD, PD or CKM) or family members’ relationship to the patient.

### Whose decision is it anyway?

The participants interviewed demonstrated a spectrum of patient-family ownership of decisions around treatment for kidney failure, though most depicted the ultimate decision as belonging to the patient. Some patients (Betty, Isabel) [pseudonyms] described securing decisional control, at times actively excluding their family members from consultations and discussions. Some patients who described having been highly independent in their treatment decision reported a history of having to “*take all responsibility*” (Betty) for domestic decisions or being “*quick to make decisions*” (Roger) in the workplace.

> *“They didn’t really get any choice in the matter. Because the choice was mine.” **Betty (85-89,F,CKM)***

Most partners described having generally agreed with the patient’s decision, and several asserted that they would agree with “*whatever decision*” (Alan’s wife) the patient had made. Meanwhile, familial influence over decisions was described, and partners, especially, were depicted as having substantial influence. For example, Jeremy’s wife was explicit about her efforts to ensure that he undertook preparation for dialysis. Indeed, where such input from partners was described, it was universally in favour of KRT:

> *“I must have been about a month persuading him to go for dialysis and in the end he says yeah, I’m not being fair to you, I’ll do it.” **Wife (75-79,F) of Jeremy (80-84,M,HD)***

Patients portrayed input from offspring (adult children or grandchildren) as being comparatively less influential. Most patients recounted how they had informed their offspring of their decision, without implication that subsequent discussions would influence their decision.

> *“I told my children, I’ve got four children, and I told them about it and see what they discovered, what they thought about it” **Clive (80-84,M,CKM)***

Some family members expressed turmoil, describing initial distress and disagreement with the patient’s consideration of CKM. Unlike partners, who appeared to have aired their views and influenced treatment plans, offspring appeared to have needed time and reflection to accept their parent’s decision to forego dialysis, and come to terms with their lack of influence in the decision:

> *“I’ve come to understand the decision. But at the time, I just felt – particularly for [Betty’s granddaughter] – are you not even going to try?” **Daughter (40-44,F) of Betty (85-89,F,CKM)***

It was infrequent for patients to describe the treatment decision as having been primarily influenced by others. Only Joan appeared to have experienced a clinician-led decision that she would “*not do dialysis*” (Joan’s daughter). A minority of participants reflected upon clinician input in decisions, typically implied to be subtle. Clinician input was depicted as having operated through the restriction of available options, including the depiction of a future “*need*” (David’s wife; Brian) for dialysis, or by highlighting the limitations of particular therapies:

> *“The only thing that was said regards that issue was…it wasn’t like a magic silver bullet that they could give that would make him back to a 30-year-old again” **Son (50-54,M) of Roger (90-94,M,CKM)***

### On death, dying and uncertain futures

For many participants, the prospect of impending kidney failure seemed to have converted an abstract awareness of mortality into concrete consideration of death and dying, often bringing distress or shock. Prognostic uncertainty was recognised by nearly all participants, and many described the challenge this brought for decision-making. Life-expectancy timeframes were rarely presented, but when mentioned, these included generous upper bounds, such as “*40 years*” (David), “*20 years*” (Clive), “*10 years*” (Jeremy’s wife), or reaching “*100*” (Roger).

> *“He basically told me that he’d been to a specialist and he had three choices and it was a bit scary because you suddenly realised that we’re all, you know, not here forever” **Daughter (60-64,F) of Clive (80-84,M,CKM)***

Throughout the interviews, dialysis was depicted as a life-prolonging treatment for kidney failure. Acceptance by patients and family members of CKM as a suitable treatment appeared to reflect, in part, their readiness to confront the proximity of the patient’s end of life. Family members’ willingness to consider this appeared related to the relationship between the patient and family member. Contrasting narratives were seen between dyads/triads where patient and family member appeared comfortable with discussions about death (Alan and his wife; Clive, his partner and daughter) and those such as Isabel and her husband, for whom such discussions appeared intolerable. Patients and family members with previous experience of bereavement tended to appear more comfortable discussing death.

> *“When I said that to my husband he shouts at me. He says he couldn’t cope without me, which puts too much pressure on me, really.” **Isabel (75-79,F,HD)***

> *“Everybody’s got to die - and I mean the older you get the more likely you are to die, that’s - I don’t think I’d be afraid to die” **Alan (85-89,M,HD)***

> *“He’s quite within his own head, quite in the knowledge that he could go tomorrow and he’s at peace in his own mind as far as I believe with that” **Wife (70-74,F) of Alan (85-89,M,HD)***

Offspring often recounted having to consider the mortality of their parent or grandparent for the first time, having not previously acknowledged “*that there’s an end coming*” (Clive’s daughter). Some elucidated a “*selfish*” (multiple participants) desire for their relative to live as long as possible, which sometimes conflicted with the patient’s willingness to forego the presumed life extension from dialysis.

> *“What’s she’s got left, she might as well enjoy…But I’m like - if you could get six months longer out of her?” **Granddaughter of Joan (85-89,F,CKM)***

Participants described disparate preparations for the future. Whilst some described future care planning, funeral planning, and financial planning for after the death, others mentioned intending to “*cross the bridge*” (Brian; Clive’s daughter) if and when future events transpired. These future planning strategies seemed at times to be used by patients to protect their autonomy and decisional influence, for example Eleanor’s advance decision to decline cardiopulmonary resuscitation rather than delegating this decision to her sons.

> *“I said to (son 1), I said if we don’t sort (son 2) out you’re going to have problems love, because he’ll want me to live forever and you’ve got to tell them that I can’t - I don’t want it. But I said it’s wrote on my notes now that I don’t want it.” **Eleanor (80-84,F,CKM)***

> *“Mum’s put ‘do not resuscitate’ on her notes and he [Eleanor’s other son] went absolutely [mad] - he said take that off.” **Son (50-54,M) of Eleanor (80-84,F,CKM)***

### Caring and being cared for

Many patients described increasing physical dependence upon their family members. Meanwhile, some patients depicted their own caring roles for their cohabiting partner or extended family. Intermingled lives and the potential impact of treatment decisions on cohabiting family members, as well as patients, was recognised.

> *“It’s now two lives are now messed about with as opposed to just one which, and I don’t want it to sound selfish, but that’s the nitty gritty.” **Son (50-54,M) of Roger (90-94,M,CKM)***

The presence of a family member who provided physical care appeared to mitigate the negative effects of decreasing functional ability. Whilst some family members described burden from this caring role, all voiced their willingness to fulfil it, because of love, a sense of reciprocity for parents who had “*done enough*” (Eleanor’s son) for them, or patient “*needs*” (Joan’s daughter). In contrast, patients voiced a reluctance to “*inflict*” (David) caring duties on their family members and a desire to avoid finding themselves in a highly dependent state (described by Clive as a “*vegetable*”):

> *“To me he’s never been a burden, to me he is not only my husband, but he is my friend and we have known each other quite a long time now and even though he can’t do the things that he used to do, I still like having him around.” **Wife (70-74,F) of Alan (85-89,M,HD)***

Being partnered in older age was seen by some participants as important for wellbeing, for example Alan, who contrasted his partnered existence with that of lonely, widowed “*old dears…[who] haven’t got a good quality of life*”. As well as being a reason for patients to favour life-prolongation, being partnered was also linked to choosing dialysis as patients appeared to factor in their partners’ preferences for this. The idea of prolonging life with dialysis to sustain a meaningful relationship with intrinsic value was also expressed:

> *“I think well we’ve been together, me and (wife) and I thought well it’s not fair to her if I just give up. So that’ll go as long as I can.” **Jeremy (80-84,M,HD)***

> *“I thought ‘well why ruin something that we’ve got?’ Whether I’m being selfish or he’s being selfish, I don’t know” **Wife (75-79,F) of Jeremy (80-84,M,HD)***

## Discussion

To our knowledge, this is the first family-level qualitative analysis focussing on the perspective of older or frailer patients and their family members approaching decisions around treatments for kidney failure, including CKM. We have shown that treatment decisions, while depicted by interviewees as belonging to patients, appear to have been shared with family members with variable levels of influence. Family members exerted this influence without necessarily having had the benefit of the education, psychosocial care and shared decision-making support offered to patients. The inextricable intertwining of patient and family members’ lives meant that both practical and existential impacts of decisions were considered. The prospect of impending kidney failure shocked many participants, and emotional distress permeated families’ considerations of treatments for kidney failure, bound up with contemplations of mortality and preferences for life-prolonging treatment. Patients who planned for dialysis with their partners appeared to have made this decision with the expectation that this would prolong the time that they would enjoy together. These expectations may prove inaccurate given uncertainty around whether dialysis substantially increases life expectancy for such patients, and the impact of dialysis on time spent with family members.^4,6^ Some of these inaccurate expectations likely result from the way treatment options are presented by clinicians.^34^ For patients considering CKM, partners appeared to exert more decisional influence compared with offspring.

The varying levels of family decision-making involvement seen in this study are in keeping with prior literature.^35–39^ Choosing dialysis to please family members may be common^40^ and has been associated with dialysis regret.^41^ Variation in familial influence (including contrasts between partners and offspring) likely reflects established decisional patterns within families, differing qualities of family relationships, and cultural norms relating to hierarchy of influence for different family members.^42–44^ Further cultural factors include the implicit expectation that patients will pre-decease their children but not necessarily their partner.^45^

An acceptance that the patient was approaching the end of their life appeared necessary for CKM to be considered as a viable treatment option for most participants, as has been seen before.^38^ Contrary to previous research,^39^ our participants did not discuss choosing CKM to shield their family members from the impact of dialysis. The tendency for family caregivers to advocate for what they perceive to be life-prolonging treatments, even where the patient favours conservative treatment, is well-recognised.^46^ This tendency is associated with factors including the level of communication between patients and family, and family caregivers’ knowledge about the disease and treatment options. Previous work suggests that adequate preparation for patients’ deaths is important, but often focusses on their own preparation, not that of family members.^47^ The ability to take part in clinical encounters and discussions over time seemed to have helped some adult children to understand their parents’ place in the life course, and rationale for favouring CKM.

The many practical impacts of treatment decisions on cohabiting family members, whose daily lives are tied up with those of patients, have been explored previously,^39^ with a recognition that the impacts can sometimes be even greater on caregivers than patients themselves.^48,49^ Those family members carrying out a physical caring role described a willingness to bear any burden, or described caring as not being a burden at all, in keeping with previous literature.^47,50^ This contrasted with the perceptions of many patients here, who seemed to struggle to adapt to being cared for. Nonetheless, the presence of a loving, dedicated family member to provide physical care seemed to mitigate the negative effects of functional impairment. This idea further advances the finding in the discrete-choice experiment which drew on this qualitative work, which showed that partnered older patients placed a relatively higher value upon living longer than upon independence, compared with their unpartnered counterparts.^51^

The strengths of this work include the novel focus upon the family unit, building upon existing research, which has mostly been non-European and focussed on the perspective of patients and clinicians,^9,10,13^ and those deciding only between KRT modalities.^9^ The dyadic approach is well-suited to exploring dynamics between patients and family members, which has been neglected by previous research. PPI involvement from conception through to analysis of this project helped to ensure the relevance of this work for patients and their family members and safeguarded ethical considerations around risk mitigation. PPI members and participants were all able to discuss mortality without high levels of distress, but one limitation is a lack of perspective from participants more distressed by existential angst that would have limited such discussions. The work is also limited by enrolment favouring the inclusion of family members sharing close relationships with patients. Consequently, findings may not apply to family members who are more distant – in geography or relationship. Patients unable to recruit family members were also excluded. Findings may also not apply to younger patients, or those for whom the benefits of KRT over CKM are less uncertain. Other weaknesses include the inherent limitation that interview studies capture participants’ accounts, which may imperfectly represent clinical encounters, and the fact that only White participants in England were recruited.

That family members can carry significant decisional influence suggests that they would benefit from education and decisional support. It is likely that patients and families will be better able to make the best decisions for them if provided with unbiased education about the potential benefits and burdens of all treatment options. Decisions about treatment for kidney failure are interwoven with considerations of mortality. As such, it would be appropriate to adopt principles of supportive and palliative care,^52^ providing physical, psychological, social and spiritual support centred on the family unit rather than the patient alone, while supporting the decision-making process. Family-centred care is well-described in paediatric^53^, palliative^52,54^, critical care^55^, mental health^56^, and disease-specific care, such as in stroke rehabilitation^57^ and dementia^58^, but is not widely described in kidney medicine, although a need has been identified.^59^

### Conclusions

In conclusion, our work asserts the central importance of family members when older patients consider treatments for kidney failure. Family members can exhibit significant decisional influence without necessarily having received decision-making support, with emotional distress from considerations of kidney failure-related mortality affecting these discussions. Intermingling of daily lives means that treatment decisions are seen to have major impacts on co-habiting family members as well as on patients. We recommend that the care of this patient group is enhanced through adoption of a family-centred approach to shared decision-making. This may help to ensure better understanding of all parties’ preferences, temper potentially negative consequences stemming from prioritisation of individualistic autonomy, and inform decisions about dialysis initiation and discontinuation in the eventuality of lost capacity. Future work should investigate the effect of this family-centred approach on decision-making processes and outcomes, as well as considering how this can be embedded in clinical practice, and how to structure our systems to facilitate this.

## Declarations

### Ethics approval and consent to participate

The study was reviewed by Surrey Research Ethics Committee and granted NHS Health Research Authority and Health and Care Research Wales approval on 2/8/18 (ref: 18/LO/1179). Reporting is in accordance with the Consolidated Criteria for Reporting Qualitative research (COREQ)^27^ (supplementary table 1). All participants gave informed, written consent for participation in the study.

### Consent for publication

All participants gave informed, written consent for their data (including quotation excerpts) to be published.

### Availability of data and materials

Raw interview data is potentially identifiable so is not made available. Please contact the corresponding author for further details.

### Authors’ contributions

LR, FJC, JC, RLM, LES and BH were involved in the original design and planning of the study. All authors were involved in the data analysis and interpretation (further detailed in methods section). RK was the major contributor in writing the manuscript but all authors contributed to the drafting and all authors read and approved the final draft.

## Acknowledgements

The authors would like to acknowledge all the research participants, those recruiting participants from all three participating centres, and the members of the patient and public involvement group for their valuable input.

## Clinical trial number

Not applicable

## List of abbreviations

CKD – chronic kidney disease; KRT – kidney replacement therapy; CKM – conservative kidney management; WHO – World Health Organisation; eGFR – estimated glomerular filtration rate; HD – haemodialysis, PD – peritoneal dialysis

---

**Supplementary figure 1:**
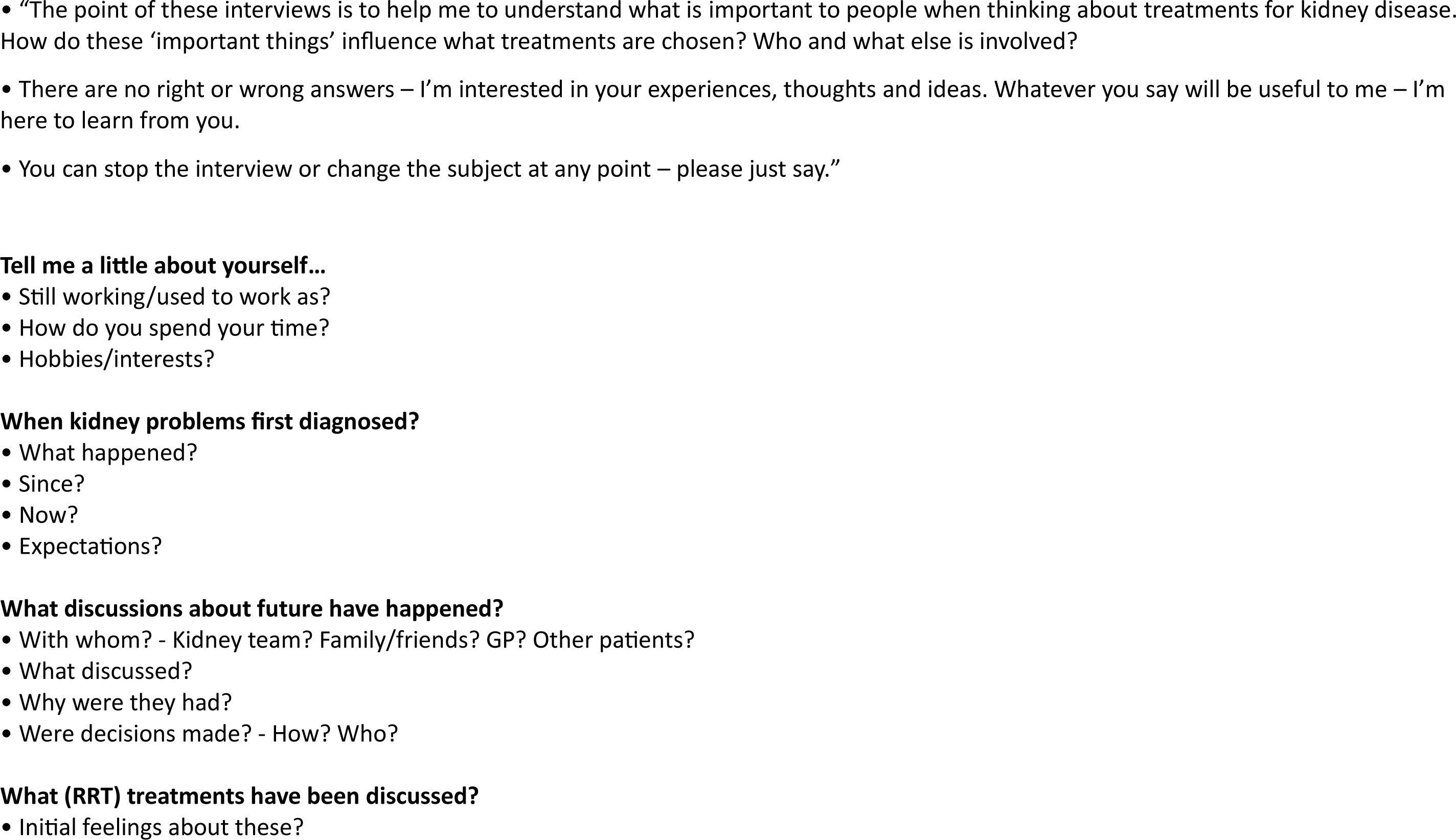
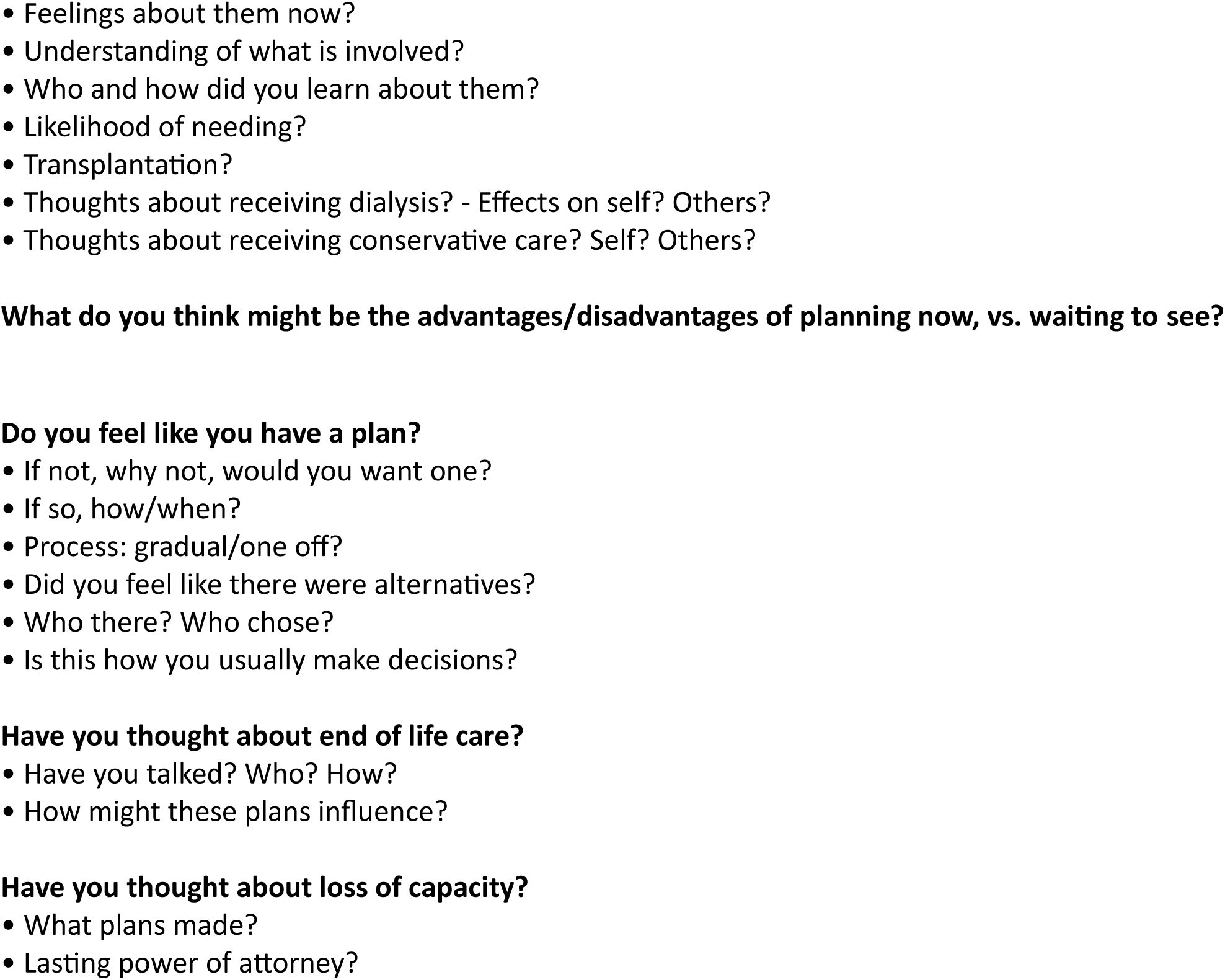
Interview topic guide.

**Supplementary figure 2:**
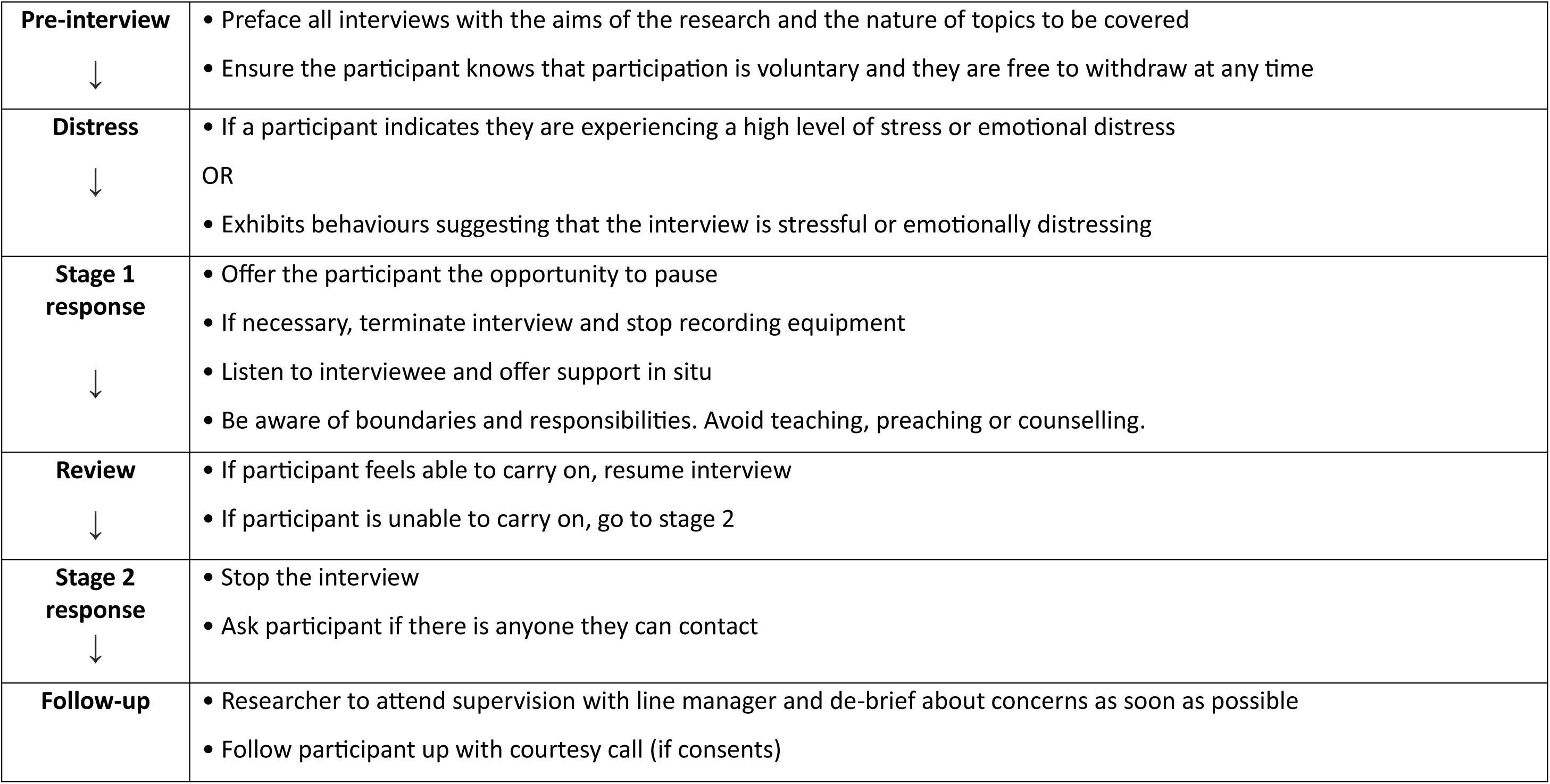
Distress protocol for qualitative interviews.

**Supplementary figure 3:**
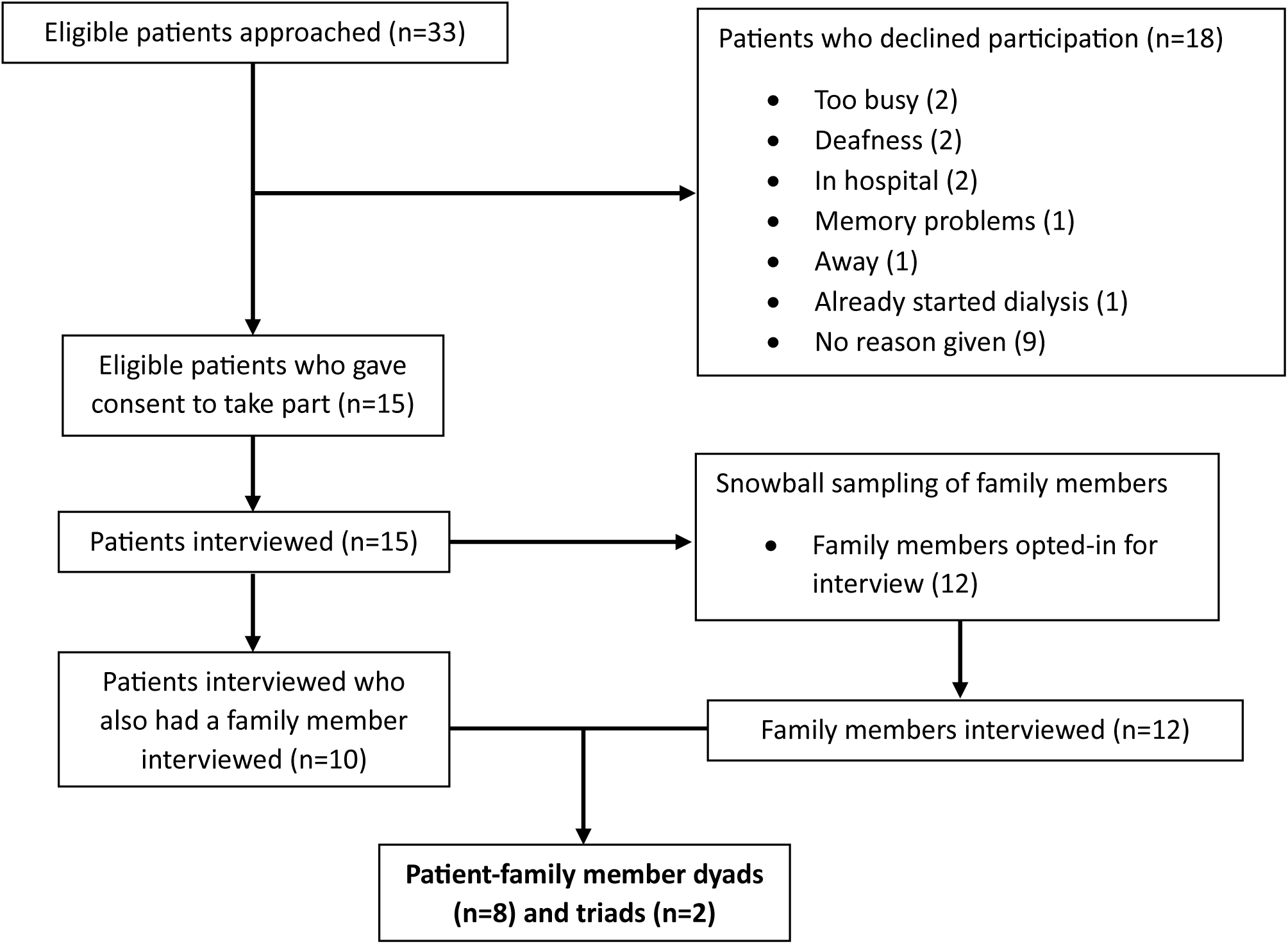
Flow diagram.

**Supplementary table 1:** Consolidated criteria for reporting qualitative studies (COREQ): 32-item checklist. ^27^.

| No. Item | Guide questions/description | Reported in section |
| --- | --- | --- |
| **Domain 1: Research team and reflexivity** |  |  |
| *Personal Characteristics* |  |  |
| 1. Interviewer/facilitator | Which author/s conducted the interview or focus group? | Materials and methods - data collection |
| 2. Credentials | What were the researcher's credentials? E.g. PhD, MD | Materials and methods - data collection |
| 3. Occupation | What was their occupation at the time of the study? | Materials and methods - data collection |
| 4. Gender | Was the researcher male or female? | Materials and methods - data collection |
| 5. Experience and training | What experience or training did the researcher have? | Materials and methods - data collection |
| *Relationship with participants* |  |  |
| 6. Relationship established | Was a relationship established prior to study commencement? | Materials and methods - data collection |
| 7. Participant knowledge of the interviewer | What did the participants know about the researcher? e.g. personal goals, reasons for doing the research | Materials and methods - data collection |
| 8. Interviewer characteristics | What characteristics were reported about the interviewer/facilitator? e.g. Bias, assumptions, reasons and interests in the research topic | Materials and methods - data collection |

| Domain 2: Study design |  |  |
| --- | --- | --- |
| *Theoretical framework* |  |  |
| 9. Methodological orientation and Theory | What methodological orientation was stated to underpin the study? | Materials and methods - analysis |
| *Participant selection* |  |  |
| 10. Sampling | How were participants selected? e.g. purposive, convenience, consecutive, snowball | Materials and methods - sampling |
| 11. Method of approach | How were participants approached? e.g. face-to-face, telephone, mail, email | Materials and methods - sampling |
| 12. Sample size | How many participants were in the study? | Results (paragraph 1) |
| 13. Non-participation | How many people refused to participate or dropped out? Reasons? | Results (paragraph 1) |
| *Setting* |  |  |
| 14. Setting of data collection | Where was the data collected? e.g. home, clinic, workplace | Materials and methods - data collection |
| 15. Presence of non-participants | Was anyone else present besides the participants and researchers? | Materials and methods - data collection |
| 16. Description of sample | What are the important characteristics of the sample? e.g. demographic data, date | Results (paragraph 2) and table 1 (participant characteristics) |
| *Data collection* |  |  |
| 17. Interview guide | Were questions, prompts, guides provided by the authors? Was it pilot tested? | Materials and methods - data collection (interview guide provided in supplementary appendix) |
| 18. Repeat interviews | Were repeat interviews carried out? If yes, how many? | Materials and methods - data collection (no repeat interviews) |
| 19. Audio/visual recording | Did the research use audio or visual recording to collect the data? | Materials and methods - data collection |
| 20. Field notes | Were field notes made during and/or after the interview or focus group? | Materials and methods - data collection |
| 21. Duration | What was the duration of the interviews or focus group? | Results (paragraph 3) |
| 22. Data saturation | Was data saturation discussed? | Materials and methods - sampling |
| 23. Transcripts returned | Were transcripts returned to participants for comment and/or correction? | No |
| **Domain 3: Analysis and findings** |  |  |
| *Data analysis* |  |  |
| 24. Number of data coders | How many data coders coded the data? | Materials and methods - analysis |
| 25. Description of the coding tree | Did authors provide a description of the coding tree? | Materials and methods - analysis |
| 26. Derivation of themes | Were themes identified in advance or derived from the data? | Materials and methods - analysis |
| 27. Software | What software, if applicable, was used to manage the data? | Materials and methods - analysis |
| 28. Participant checking | Did participants provide feedback on the findings? | No |
| *Reporting* |  |  |
| 29. Quotations presented | Were participant quotations presented to illustrate the themes/findings?<br>Was each quotation identified? e.g. participant number | Results (under thematic headings) and table 2 (illustrative quotations) |
| 30. Data and findings consistent | Was there consistency between the data presented and the findings? | Results (under thematic headings) and table 2 (illustrative quotations) |
| 31. Clarity of major themes | Were major themes clearly presented in the findings? | Results (under thematic headings) |
| 32. Clarity of minor themes | Is there a description of diverse cases or discussion of minor themes? | Table 2 (illustrative quotations - subthemes) |

